# PHOTONAI-Graph - A Python Toolbox for Graph Machine Learning

**DOI:** 10.1101/2023.06.22.23291748

**Authors:** Jan Ernsting, Vincent Holstein, Nils R. Winter, Kelvin Sarink, Ramona Leenings, Marius Gruber, Jonathan Repple, Benjamin Risse, Udo Dannlowski, Tim Hahn

## Abstract

Graph data is an omnipresent way to represent information in machine learning. Especially, in neuroscience research, data from Diffusion-Tensor Imaging (DTI) and functional Magnetic Resonance Imaging (fMRI) is commonly represented as graphs. Exploiting the graph structure of these modalities using graph-specific machine learning applications is currently hampered by the lack of easy-to-use software. PHOTONAI Graph aims to close the gap between domain experts of machine learning, graph experts and neuroscientists. Leveraging the rapid machine learning model development features of the Python machine learning API PHOTONAI, PHOTONAI Graph enables the design, optimization, and evaluation of reliable graph machine learning models for practitioners. As such, it provides easy access to custom graph machine learning pipelines including, hyperparameter optimization and algorithm evaluation ensuring reproducibility and valid performance estimates. Integrating established algorithms such as graph neural networks, graph embeddings and graph kernels, it allows researchers without significant coding experience to build and optimize complex graph machine learning models within a few lines of code. We showcase the versatility of this toolbox by building pipelines for both resting–state fMRI and DTI data in the hope that it will increase the adoption of graph-specific machine learning algorithms in neuroscience research.

## Introduction

Graph data is ubiquitous throughout biomedical research and can be found in many different fields. In neuroscience, graph representations are of particular interest as the neuronal connections within the brain are a naturally occurring graph structure. These graphs arise both on the microscopic level in cellular connection networks and the macroscopic level such as higher-order brain circuits. Functional connectivity and diffusion tensor imaging (DTI) are the two most commonly used modalities for studying these circuits in vivo. The two modalities allow for the construction of brain connectivity graphs, which can then be studied using graph theoretical approaches. Different established toolboxes allow neuroscientists to apply classical graph theoretical approaches using statistical analysis. However, multivariate graph analyses such as graph machine learning pipelines are mostly limited to a domain expert group, limiting the accessibility for many neuroscientists.

In classical graph analysis graph properties are calculated, sometimes referred to as graph measures, which are then analyzed using statistical analysis tools such as general linear models (GLMs). This approach has been increasingly adopted in neuroimaging, which is in part due to the availability of toolboxes, that support these analyses [1, 2]. These toolboxes are usually geared towards neuroimaging often specializing in one type of connectivity modality. One popular toolbox is Brain Connectivity Tools which offers a graphical user interface and implements the most important graph analysis methods based on graph measures [3]. Other frequently used toolboxes for analyzing brain connectivity graphs using graph measures include but are not limited to, Network-based Statistics, eConnectome, CONN, GAT, GTG, BASCO, GRETNA or BRAPH [4–11].

These neuroimaging-specific toolboxes support basic graph analysis, but not graph machine learning analyses. As one of the first toolboxes, the GraphVar 2.0 toolbox expands this framework to the area of machine learning by allowing users to perform machine learning on extracted measures [12]. This represents a novel approach towards creating low-dimensional graph representations that capture important information about the inherent graph structure which are then used for machine learning. However, this represents only a small area of the field of graph machine learning which has exponentially grown in recent years.

The approach of creating lower-dimensional graph representations is at the core of the field of graph machine learning. In this new machine learning subfield the main directions of research are graph embeddings, graph kernels and graph neural networks [13]. Graph embeddings project to a lower dimensional representation that leverages graph information and makes it accessible in Euclidean space [14]. Graph kernels are functions that either extract feature representations or calculate similarity measures between graphs, which can be leveraged using kernel methods [15]. Significant overlap exists between the two fields of research. Lastly, there are graph neural networks, which are a form of neural networks that leverage graph information, by performing neural message passing. Here information flows between nodes and is updated using neural network functions [16, 17].

In neuroimaging, most research uses graph measures to predict outcomes of interest. Recently researchers have begun to use graph machine learning in neuroimaging and even built graph neural networks specifically targeting brain connectivity matrices [18, 19]. These approaches however are still much less common than classic graph analysis, partly because they require require significant technical knowledge. While the neuroimaging community has developed packages for graph measure analysis on brain-derived graphs, the graph machine learning community has developed specialized packages for graph machine learning. However, these require advanced coding experience and usually cover only one particular (sub-)field. Examples include *gem* for graph embeddings, *grakel* for graph kernels and deep graph library (*dgl*) for graph neural networks [20–23]. All of these toolboxes require programming expertise and technical knowledge for their usage, which bars many scientists without such background from routinely using these for their analyses. This causes an accessibility gap for many neuroscience researchers, which is not addressed by any of these toolboxes. Therefore there is currently no toolbox that implements a wide array of graph machine algorithms in an accessible manner. Existing toolboxes in neuroimaging focus on classical graph analysis or graph measure-derived machine learning, while graph machine learning toolboxes focus on one type of algorithm and require significant computational expertise. Among the computational challenges are the incompatibility of different graph libraries, varying data structures and error-proneness of such conversions. To address this gap we developed our graph machine learning toolbox called *“PHOTONAI Graph”*.

The integration of this toolbox into the *PHOTONAI* package provides increased accessibility of complex algorithms via pre-defined keywords, extended pipeline functionality such as stacking operations, a high degree of automation with variable hyperparameter optimization and cross-validation strategies, easy visualization and model sharing options ([24]). This provides neuroscientists with access to best practices in machine learning and rapid prototyping capabilities, which will strongly improve model building without the need to learn and code a large body of graph algorithms.

## Methods & Software

*PHOTONAI-Graph* is a *scikit-learn*-compatible *Python* package that is an extension to the PHOTONAI toolbox [24], which provides a high degree of automation of the repetitive steps during model development. As an extension to the *PHOTONAI* framework, it uses the functionality of the *PHOTONAI* toolbox in the context of graph machine learning. This allows an out-of-the-box usage of graph algorithms with fully automated hyperparameter optimization, model evaluation and algorithm selection procedures. This integration also enables seamless access to *PHOTONAI’s* convenience features such as result visualization and model sharing and integrates within *PHOTONAI’s* structured and easy-to-learn syntax, condensing machine learning analysis to a few lines of code.

The *PHOTONAI-Graph* toolbox consists of multiple modules which correspond to different types of data transformations, learning algorithms, and utilities. In the following sections, we explain each of the different toolbox modules, and our testing strategy to ensure functionality, public access, documentation and software dependencies.

### Organisation

The toolbox is designed to adapt to various types of graph data, with a focus on connectivity data. It is modality-agnostic however, as graph machine learning pipelines can be built for other types of graph data, as long as it is imported in a compatible format. For various types of graph data, the toolbox supports the development of whole graph classification or regression pipelines.

Pipeline development is supported by 8 main modules: *Graph Constructors, Graph Measures, Graph Kernels, Graph Embeddings, Graph Neural Networks, Controllability, Graph Conversions* and *Graph Utilities*. These modules cover key areas of current graph machine learning research and allow for the use of state-of-the-art graph machine learning in a few lines of code.

Neuroscientific graph data, such as connectivity matrices, can be transformed using *Graph Constructors*, to ensure adequate graph representation via adjacency matrices. These can be further transformed with graph-specific dimensionality reduction techniques using *Graph Kernels, Graph Embeddings, Graph Measures* or *Controllability* functions. The transformed data can then be passed to an estimator. Alternatively, graph properties can directly be estimated using a *Graph Neural Network*.

Conversions between different graph formats are handled by the *Graph Conversions* module, while the *Graph Utilities* module supplies different utilities such as drawing functions. Each module will be described in brief below.

### Graph Constructors

The Graph construction module implements graph construction techniques that transform connectivity matrices into adjacency matrices to reduce noise and complexity, speed up computation and increase the signal-to-noise ratio. This transformation is especially important when working with highly dense data (i.e. fully connected graphs) such as resting-state connectivity.

Implemented techniques include thresholding based on cutoff values, the percentile rank, a window of cutoff values or percentile ranks, kNN-based adjacency formation, spatial proximity-based adjacency formation and random walks on kNN graphs. These transformations are applied individually to each graph. It is also possible to use the *PopulationAveragingTransform* class to build an average graph across each fold as described by Ktena et al. [25].

Users can also decide to apply different encodings for node features, such as a one-hot encoding, similar to indicator variable encodings or they can use the unfiltered connectivity matrix as a feature matrix. This way the feature matrix, which can later be used to define node and edge features, contains all the original information before any transformation was performed.

**Fig. 1.**
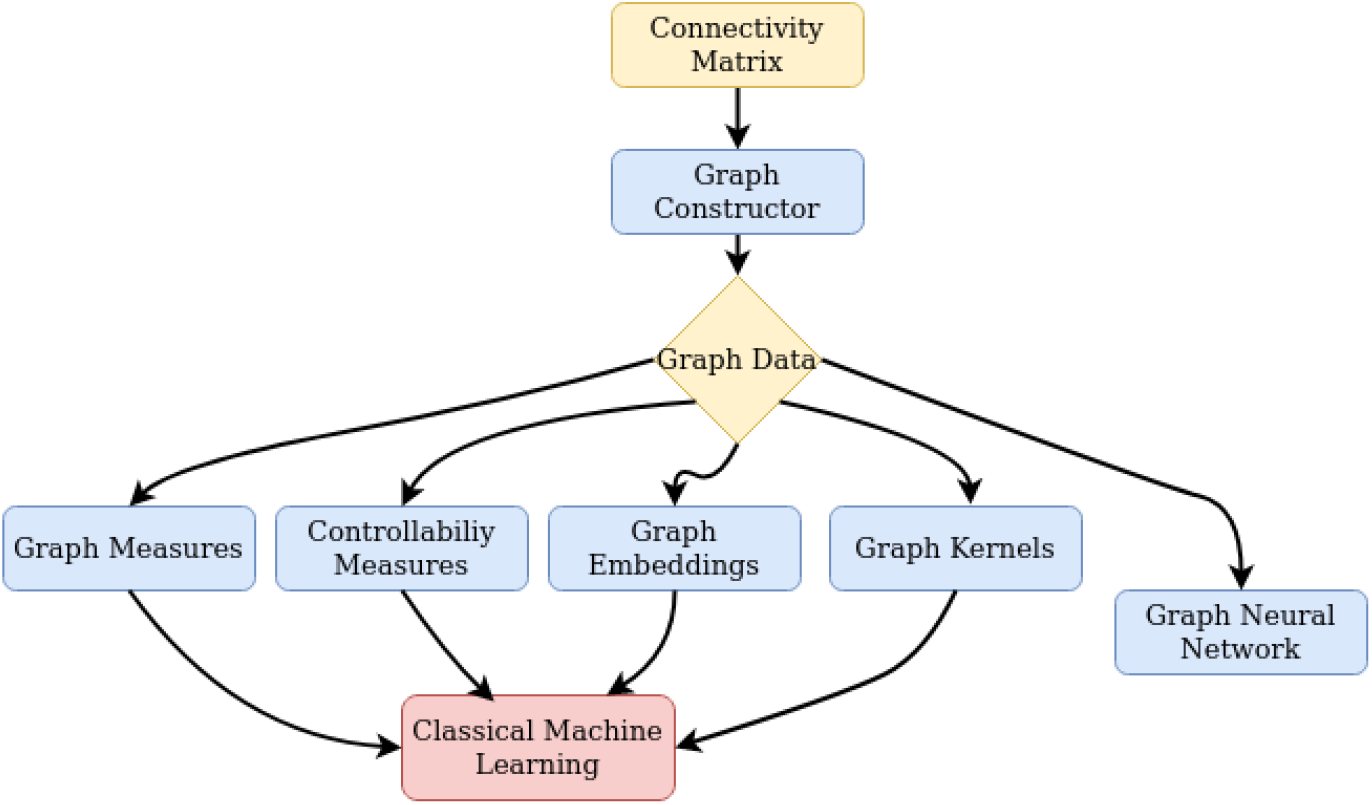
Modules of PHOTONAI-Graph. The PHOTONAI-Graph package features multiple modules for transforming and predicting from graph data. Starting from connectivity matrices, the graph constructor module allows for graph construction with a range of established and novel thresholding and edge selection strategies. The derived graph data can be used by either transforming it with graph measures, network controllability measures, graph embeddings or graph kernels to acquire graph property-preserving representations that can then be used in concordance with classical machine learning estimators. Alternatively, users can directly use the graph data by feeding it to a graph neural network.

### Graph Measures

The Graph Measure Transform module implements the *NetworkxMeasureTransform* and *IgraphMeasureTransform* class which calculate graph measures based on the *networkx* or *igraph python* package and concatenates them into a feature vector [26, 27]. Whole graph, node and edge measures can be combined to provide a low-dimensional feature representation of the graph. These feature vectors can then be analyzed using machine learning estimators. The choice of desired graph measures is a hyperparameter during model development.

To reduce runtime these calculations can be parallelized based on *tqdm* [28]. The *GraphMeasureTransform* class can also be used to extract graph measures into a CSV file for further statistical analysis outside of graph machine learning pipelines.

### Graph Embeddings

The graph embedding module implements graph embeddings, based on a modified version of the *gem python* package [21]. These include the Asymmetric Transitivity Preserving Graph Embedding, Laplacian Eigenmaps and a locally linear embedding. As graph embeddings are not calculated between graphs but are a direct low-dimensional representation of the graph, they do not require the use of kernel methods.

To ensure the correct functioning of the *gem* software and avoid dependency conflicts we have developed a modified version of the *gem python* package which is publicly available (https://github.com/jernsting/nxt_gem) for this project.

### Graph Kernels

The Graph Kernel module implements Graph Kernels based on the *grakel python* package, allowing for the usage of kernels currently available in *grakel* [22]. All available *grakel* kernels can be optimized with respect to their hyperparameters via the *GrakelTransformer* class. These kernels can then be used in a pipeline in combination with kernel methods such as support vector machines.

To ensure the correct conversion of input graph data into the required *grakel* format as part of a machine learning pipeline the module also contains the *GrakelAdapter* class, a transformer that handles these conversions. This adapter also allows the user to select different feature construction options if the graphs are converted from connectivity matrices.

### Graph Neural Networks

The graph neural networks module currently implements six different Graph Neural Network (GNN) architectures, based on the deep graph library (*dgl*) *python* package and the *pytorch* library [23, 29]. These GNNs allow the user to use different architectures for both whole graph classification and regression tasks. As GNNs combine both graph representation learning and feature estimation, they do not require an additional estimator in the pipeline.

The GNN module consists of a *dgl_base* class that handles training and conversion steps shared between the different architectures, a set of graph neural network-specific utilities and architecture wrappers for graph convolutional networks, simple graph convolutional networks and graph attention networks. Each GNN architecture allows for wide-ranging hyperparameter optimization including layer size, depth, learning rate and the number of training epochs.

### Controllability

The controllability module implements the modal and average controllability on graphs which can be used as a low-dimensional representation of the graph. These function are adapted from *nctpy* ([30–32] (https://github.com/BassettLab/nctpy). These lowdimensional representations can then be combined with estimators for regression or classification tasks. They can also be used outside of graph pipelines to calculate the modal and average controllability for further statistical analysis.

### Graph Conversions

Graph Conversions are a collection of different conversion functions that handle conversions between different graph formats, namely: *networkx* graphs, *numpy* arrays, *scipy* sparse arrays, *dgl* graphs and *grakel* graphs. This facilitates the data flow between the available modules. It also implements the *check_dgl* function that ensures that incoming data is converted into the *dgl* format for graph neural networks.

### Graph Utilities

The graph utilities module is a collection of helper functions that allow for the plotting of graphs in different formats, checking certain properties on graphs, fisher- and z-transformations and the generation of random data. The plotting functions allow the user to visually inspect the input data, both for connectivity matrices and *networkx* graphs.

### Toolbox documentation and installation

The entire software is made publicly accessible as a repository on GitHub (https://github.com/wwu-mmll/photonai_graph) and is published under an MIT License. To facilitate public use and allow easier use by researchers without a computer science background, we created a documentation website hosted on GitHub Pages (https://wwummll.github.io/photonai_graph/). Here we give a detailed description of the different classes and functions, along with their parameters and arguments. Current installation instructions can also be found on the website or in the GitHub repository.

To ensure the correct functioning of our code the software was unit tested with >90% coverage. Additionally, scenario tests ensure the correct integration of the different modules.

### Analysis Data

For all analysis demonstrations, we used two publicly available imaging datasets. 10Kin1Day and the Human Connectome Project Young Adult (HCP-YA) dataset [33, 34].

The 10KIn1Day dataset contains >8000 participants processed with the Cammoun Desikan-Killiany atlas parcellation containing 128 regions resulting in 16384 edges [33]. For each participant five measures of connection strength are available as edge weights: 1) Number of streamlines (NOS), 2) average fractional anisotropy (FA), 3) average mean diffusivity (MD), 4) average length of reconstructed streamlines and 5) streamline density. We used all 8163 participants (4339 male, 3824 female) for which sex information and DTI matrices with number of streamlines was available in our analysis. Further information can be found in van den Heuvel et al, 2019 [].

The HCP-YA dataset is a dataset of >1000 young adults with resting-state fMRI (rs-fMRI) and preprocessed parcellations available [34]. Resting-state fMRI scans were preprocessed running a spatial ICA with 15, 25, 50, 100, 200 and 300 numbers of components from FSL’s MELODIC tool. Network matrices were derived using the FSLNets toolbox on the node time series. We selected all preprocessed matrices with 50 components and available sex or strength information for our analysis resulting in 1003 individuals being included in sex prediction and 1002 individuals being included in our grip strength regression pipeline. The sex analysis contained 469 males and 534 females. Grip strength was measured by the NIH Toolbox Grip Strength Test using dynamometry and is provided as part of the HCP-YA data release. It is measured in pounds and adjusted for age with a sample range of 45.41 to 154.59.

All analysis scripts are available on GitHub (https://github.com/wwu-mmll/photonai_graph_usecases/) in a dedicated repository with data handling scripts for convenient handling of the datasets.

## Results

To demonstrate the versatility of our toolbox, we conducted three different analyses on two publicly available datasets to show both the versatility of the toolbox and potential use cases for it. It is important to note that for each of these demonstrations, we aim not at constructing the best available model, but rather showcase the versatility of our toolbox and the potential analyses that could be conducted with it.

### Predicting Sex from DTI

First, we build a sex classification model on the 10KIn1Day dataset to show the application of the toolbox to DTI data. The pipeline used a threshold constructor with a fixed threshold of 0.1, a stack of controllability transformation with average and modal controllability and a Support Vector Classifier as a predictor. Optimizable hyperparameters were the number of components in the PCA, the C parameters and the kernel of SVM. The C parameter was allowed to vary between 0.1, 1, 2.5 or 10. The potential kernels were either linear or rbf. We used 10x10 K-fold nested crossvalidation with random grid search and 50 configurations for hyperparameter selection selecting configurations based on accuracy. We also calculated balanced accuracy, recall and precision.

**Table 1.** Predictive performance for sex classification of the best pipeline trained on the 10KIn1Day DTI dataset.

| Metric | Test Mean ± Standard Deviation |
| --- | --- |
| Accuracy | 0.642 ± 0.024 |
| Balanced Accuracy | 0.623 ± 0.031 |
| Precision | 0.617 ± 0.101 |
| Recall | 0.511 ± 0.151 |

The overall best pipeline configuration was able to classify sex with an accuracy of 0.64, balanced accuracy of 0.62, a recall of 0.51 and a precision of 0.61. It used 100 PCA components, a C parameter of 1 and an rbf kernel.

### A. Predicting Sex from resting-state fMRI

Next, we build a sex classification model from the HCP-YA dataset to show an application to fMRI data. The pipeline consisted of a percentage constructor with a threshold of either 50, 75 or 90 and a Graph Convolutional Neural Net Classifier with 1 to 3 hidden convolutional layers, hidden dimensions of 32, 64, 128 or 256 and training epochs of either 50, 100, 250 or 500. We used a 10x10-fold nested cross-validation for hyperparameter optimization with the best configuration being selected based on accuracy. We used a Bayesian hyperparameter optimization strategy provided by *sk_opt* with the lower confidence bound as the acquisition strategy and a quasirandom Sobol vector point generator with 15 initial points and 25 configurations. We again calculated balanced accuracy, recall and precision.

**Fig. 2.**
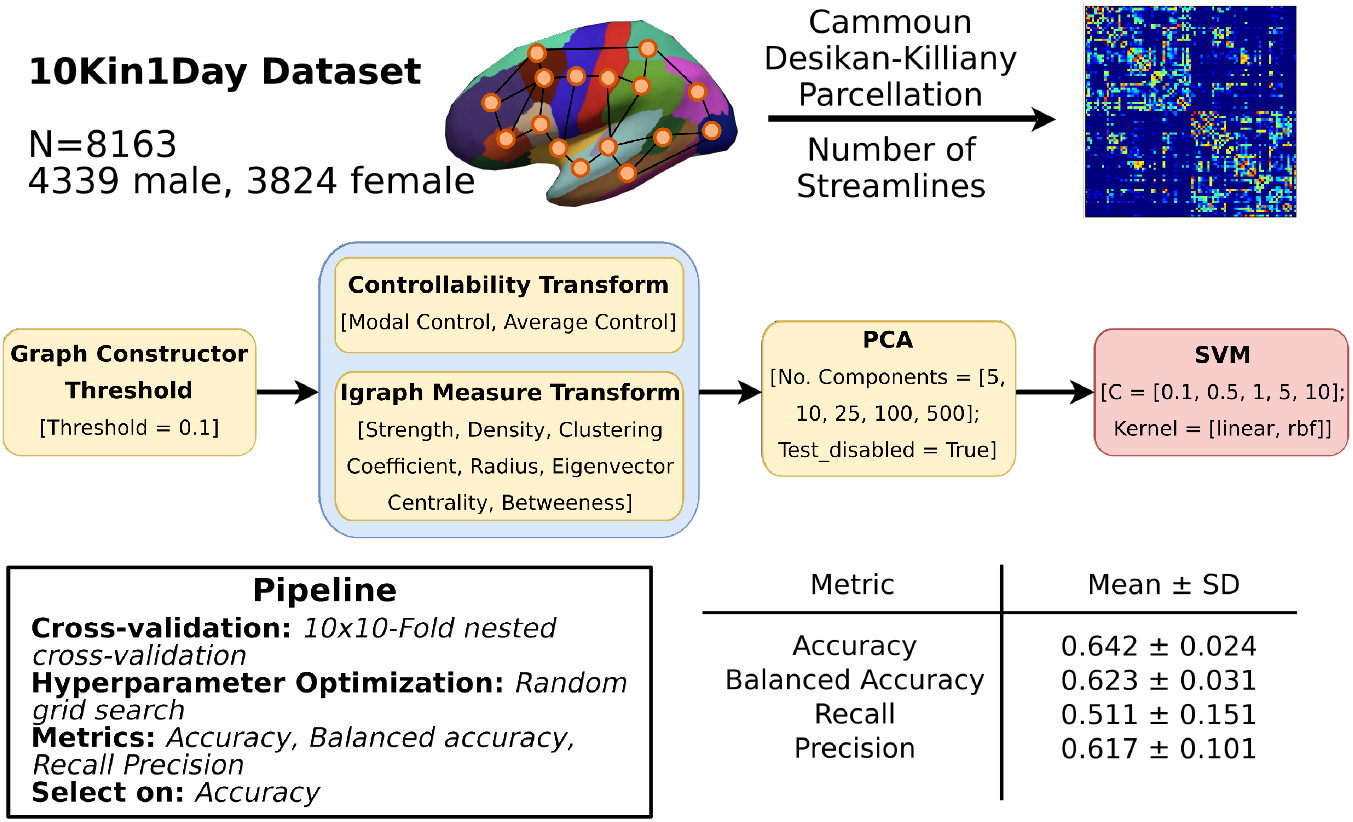
Predicting sex from DTI. We built a sex classification model from the 10KIn1Day dataset (N=8163; 4339 male, 3824) using connectivity matrices with the number of streamlines derived from the Cammoun Desikan Killiany parcellation to showcase a pipeline for DTI derived connectivity. The pipeline consisted of threshold graph constructor, a stack of a controllability and igraph measure transform, a PCA and an SVM as an estimator. The best model was selected using a 10x10 K-Fold nested cross-validation with the best model selected on accuracy. The best pipeline configuration classified sex with an accuracy of 0.64, a balanced accuracy of 0.62, recall of 0.51 and a precision of 0.61. The model used 100 PCA components, a C parameter of 1 and an rbf kernel.

The overall best pipeline configuration was able to classify sex with an accuracy of 0.74, balanced accuracy of 0.74, precision of 0.72 and recall of 0.74. The selected percentage threshold was 90, while the Graph Convolutional Classifier with the best performance used 50 training epochs, 2 hidden layers, and a hidden layer size of 256.

**Table 2.** Predictive performance for sex classification of the best pipeline configuration trained on the HCP-YA dataset.

| Metric | Test Mean ± Standard Deviation |
| --- | --- |
| Accuracy | 0.749 ± 0.026 |
| Balanced Accuracy | 0.747 ± 0.026 |
| Precision | 0.727 ± 0.035 |
| Recall | 0.740 ± 0.053 |

### Predicting grip strength from resting-state fMRI

Lastly, we build a strength prediction model to showcase the building of a regression model using the HCP-YA dataset to showcase the application of our toolbox to a regression problem.

We constructed a pipeline consisting of a threshold constructor with a threshold between 0 and 1, and a Graph Attention Neural Net Classifier with 1 to 3 hidden convolutional layers, hidden dimensions of 32, 64, 128 or 256 and training epochs of either 50, 100, 250 or 500. We used 10x10-fold nested cross-validation for hyperparameter optimization with the best configuration being selected based on Pearson correlation. We also calculated mean absolute error and explained variance. For hyperparameter optimization, we used the same *sk_opt* strategy as previously in the resting-state fMRI sex prediction.

The overall best pipeline configuration achieved a Pearson correlation of 0.36, a mean absolute error of 16.58 and a variance explained of 0.135. The threshold value for the constructor was 0.877, and the Graph Convolutional Net Regressor used 250 training epochs with 3 hidden layers and a hidden layer size of 32.

**Table 3.** Predictive performance for grip strength prediction of the best pipeline configuration trained on the HCP-YA dataset.

| Metric | Test Mean ± Standard Deviation |
| --- | --- |
| Pearson Correlation | 0.361 ± 0.068 |
| Mean Absolute Error | 16.584 ± 0.814 |
| Variance Explained | 0.135 ± 0.049 |

## Discussion

The previous sections showcase how our toolbox can be applied to different types of imaging-derived connectivity data to perform graph machine learning within a few lines of code. Having established the utility of the tool, we discuss the utility and value of this toolbox within the framework of existing toolboxes for different types of graph analyses.

### A new toolbox for graph machine learning

Graph machine learning is an important subfield of machine learning, due to the common occurrence of graph data [35]. Yet, despite their ubiquitous occurrence, machine learning that leverages the information by graph structures has only recently come into focus. While packages for specific types of graph machine learning exist (e.g. gem for graph embeddings), no package makes it possible to use these various approaches in an easy end-to-end framework. This is where *PHOTONAI-Graph* fills a gap.

Through integration with *PHOTONAI* the *PHOTONAI-Graph* toolbox allows scientists to set up graph complicated machine learning pipelines with minimal coding expertise, providing access to automated pipeline selection, best practices such as nested cross-validation, advanced hyperparameter optimization strategies and model visualization. This allows researchers without strong coding expertise to use graph machine learning with correct algorithm evaluation. Furthermore, it significantly decreases development time, allowing for rapid prototyping while safeguarding against conceptual errors such as information leakage between, training, validation and test folds.

**Fig. 3.**
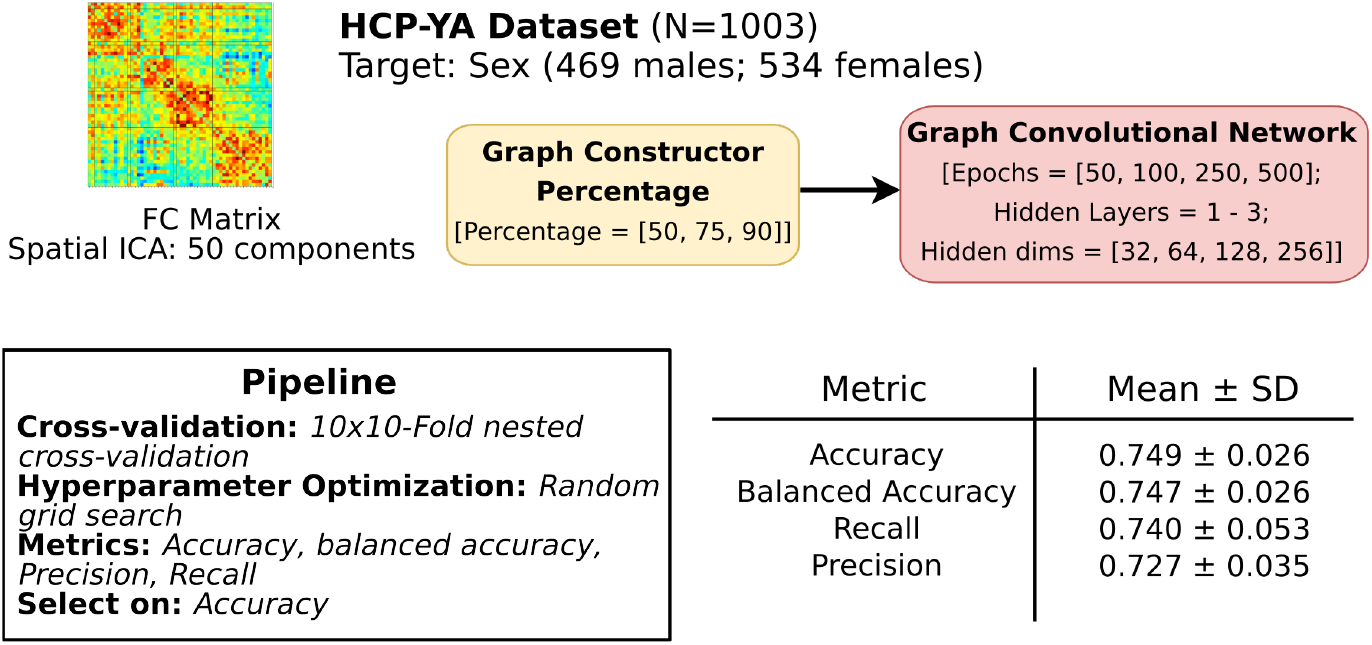
Predicting sex from fMRI. We build a sex classification model on the HCP-YA dataset (N=1003; 469 males, 534 females) using functional connectivity matrices based on an spatial ICA with 50 components to showcase an application with functional connectivity data. The pipeline contained a percentage constructor and a Graph Convolutional Neural Net Classifier. The best model was selected based on accuracy within a 10x10 K-fold nested cross-validation using a bayesian hyperparameter optimization strategy. The best model classified sex with an accuracy of 0.74, balanced accuracy of 0.74, precision of 0.72 and recall of 0.74 with percentage threshold of 90 and a Graph Convolutional Classifier with 50 training epochs, 2 hidden layers, and a hidden layer size of 256.

**Fig. 4.**
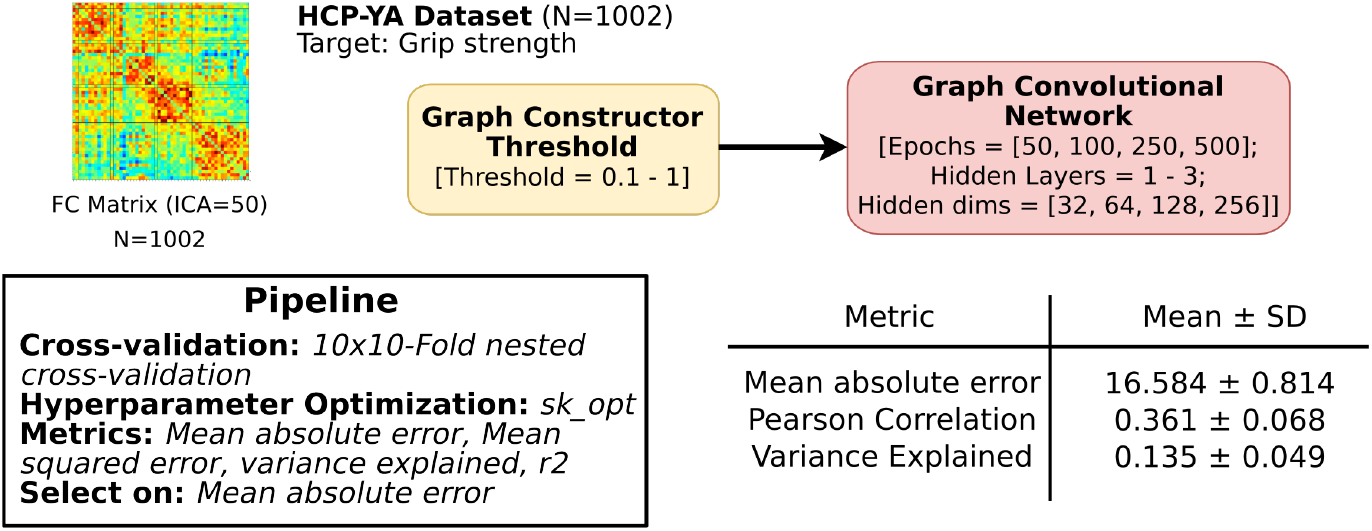
Predicting grip strength from fMRI. We built a grip strength regression model on the HCP-YA dataset (N=1002) using functional connectivity matrices derived using a spatial ICA with 50 components to showcase an application to regression data. The pipeline contained a threshold constructor and a Graph Convolutional Network Classifier. The best model was selected on mean absolute error with a bayesian hyperparameter optimization strategy within a 10x10 K-Fold cross-validation. The best model predicted grip strength with a mean absolute error of 15.58, a pearson correlation of 0.36 and a variance explained of 0.13 using a threshold value of 0.877 and a Graph Convolutional Net Regressor with 250 training epochs, hidden layers and a hidden layer size of 32.

### Software for graph machine learning

Major fields in graph machine learning are graph embeddings, graph kernels and graph neural networks [13, 36]. Network controllability and graph analysis are not part of classical graph machine learning but can be used to reduce dimensionality while preserving graph information. Different packages exist for these approaches, many of which have been integrated into the *PHOTONAI-Graph* toolbox.

The gem package is a *python* package that implements graph embeddings. It covers static graph embeddings and dynamic graph embeddings are currently under development [21]. It is to our knowledge the only package that specifically covers graph embeddings in *python*. By integrating *gem* into the *PHOTONAI-Graph* toolbox, we provide users with easy access to graph embedding techniques and make these embeddings available for *scikit-learn*-based machine learning pipelines.

For graph kernels, the *grakel* package has been developed as a *python* package that implements 18 different graph kernels as *scikit-learn*-compatible transformer classes [22]. While the *scikit-learn*-based API *grakel* kernels can easily be used in machine learning pipelines, *grakel* requires a specific *grakel* graph object, which requires additional coding for format conversion. Integrating *grakel* into our toolbox allows for automatic graph conversion and hyperparameter optimization as part of machine learning pipelines, which lowers the entry bar for graph kernel-based analyses. Alternative packages for graph kernels such as *graphkernels* [37] or *graphkit-learn* [38] were not selected for integration as *grakel* provides the largest amount of available graph kernels with an active community and regular package maintenance.

For graph neural networks different packages exist, mostly written in *python*. Among the most widely used packages are *stellargraph, spektral, Pytorch Geometric, graphnets* and *dgl* [23, 39–41]. All of these packages provide frameworks for the creation of graph neural networks, with different neural network dependencies. Of these, *dgl* provides the highest amount of flexibility as it is backend-agnostic and supports *PyTorch, TensorFlow* and *MxNet* [23, 29, 42, 43]. It is regularly updated and supported by an active developer community. *dgl* models in *PHOTONAI-Graph* use a *pytorch* back-end. As graph neural networks rapidly develop, *dgl* allows for an easy switch to another backend in the future.

Next to graph machine learning, classical graph analysis has gained increasing popularity in the area of neuroimaging and this is reflected by a range of toolboxes that offer the extraction of different graph measures from DTI and rs-fMRI data. Notable examples include GAT, Brain Connectivity Toolbox, GRETNA, Dynamic Graph Metrics and NeuroPycon [3, 10, 44–46]. All these toolboxes implement the extraction of graph measures from brain connectivity networks and often provide full pipelines for brain connectivity analyses. These toolboxes however are focused on brain connectivity and are not written to be applied to other forms of graph data. For this reason, we chose the *networkx* and *igraph python* package for our graph *GraphMeasureTransform* module which is agnostic to the data source as long as graphs can be transformed into the *networkx* or *igraph* graph format. This allows us to adapt the toolbox to other types of graph data, outside of neuroimaging. Providing both graph measure transformations, users are able to choose between *networkx* for breadth and *igraph* for speed when needed.

Network control theory has recently been introduced to brain connectivity networks [30]. As network controllability measures relate to psychiatric disease we have included two established measures of brain controllability in our controllability module [47–49]. Here we use our own implementation of average and modal controllability based on Tang et al [50]. As research in this area expands and network controllability toolboxes emerge, we plan to include these as part of *PHOTONAI-Graph*.

### Software for graph machine learning in neuroimaging

In the area of neuroimaging, a python package that incorporates existing neuroimaging toolboxes into a unified framework is the NeuroPyCon package [46]. NeuropyCon combines various toolboxes that allow connectivity-based analyses and incorporates them into a single python framework, which is an extension of NiPype [51]. It includes shareable parameters to facilitate reproduction in neuroimaging pipelines and can work with source data from fMRI, MEG and EEG. It specializes in graph analysis using the tools of existing toolboxes like CONN or Network-Based Statistics (NBS) but does not cover graph machine learning. As our toolbox does not focus on deriving graph measures but on graph machine learning it does not stand in competition with existing graph analysis toolboxes for neuroimaging.

A toolbox that specializes in graph-specific machine learning is *GraphVar 2*.*0*. It is a neuroimaging-specific toolbox, that addresses machine learning based on graphs using graph measures. It is based on MATLAB and is extended to SPM, to allow easy integration with existing machine learning tools. Like *PHOTONAI-Graph*, it allows for nested cross-validation, hyperparameter optimization, model selection and evaluation [12]. It does not support model sharing however and machine learning methods are limited to extracting graph measures in order to perform classical machine learning analyses. Furthermore, it is not agnostic to the type of input data, as it only supports brain connectivity analyses. Here *PHOTONAI-Graph* widely expands the space of usable algorithms, covering state-of-the-art graph machine learning techniques from multiple subfields.

### Limitations and future developments

One key limitation of *PHOTONAI-Graph* is that despite strongly increasing accessibility of state-of-the-art graph machine learning techniques it still requires a certain degree of technical knowledge for preprocessing and preparation of the input data. It is not constructed as a front-to-end pipeline tool, which means that technical knowledge is still required for the construction of connectivity matrices. This can be done using established tools such as CONN, NeuroPycon or CATO which sometimes offer GUI support [6, 46, 52].

It also requires basic familiarity with *numpy* array notation, for input data to have the required form: Individuals x nodes x nodes x matrix types (adjacency, features). GUI support is currently not available and basic coding skills are required for setting up pipelines. We plan to introduce *PHOTONAI-Graph* to the PHOTONAI Wizard (https://photon-ai.com/wizard) in the near future to allow users to construct *PHOTONAI-Graph* pipelines using a GUI. Another limitation is the focus on brain connectivity matrices. While it is possible to perform other types of graph analyses with *PHOTONAI-Graph*, the input data requires conversion into an appropriate format. This requires a higher degree of technical knowledge. For future updates, we plan to expand the number of supported data structures, expand import and conversion functions, include sparse support and develop an internal data structure that can accommodate large and sparse graph structures.

Furthermore, we plan to include support node classification and link prediction pipelines in the future. These are two vital areas of graph machine learning, which can also be applied to brain connectivity graphs. While most applications of graph machine learning to brain connectivity focus on whole graph prediction, the application of methods to brain graphs could enable researchers to answer new questions about brain connectivity. We plan to introduce these changes along with sparse support for giant single graphs.

## Conclusion

Our toolbox incorporates various graph machine learning packages and makes their algorithms available via a *scikit-learn*-compatible API that could be used with the *PHOTONAI* package or outside of it. It offers a unified framework for graph machine learning, without the need for manually converting graph data from one format to the other. This eases model building and hyperparameter optimization, allowing researchers from different biomedical research fields to use graph machine learning for their own analyses. Furthermore, the toolbox delivers a set of functions that can be specifically used with connectivity matrices, such as the graph constructor classes. They include established and novel approaches geared toward machine learning on neuroimaging data [53]. Through integration with the *PHOTONAI* environment, it also offers the functionality provided by the *PHOTONAI* toolbox, like model sharing, pipeline evaluation and model interpretation.

In conclusion we present a novel toolbox that allows neuroimaging researchers to construct graph machine learning pipelines within a few lines of code. With this toolbox, researchers can now easily apply graph-based machine learning methods to their data and extract valuable insights that were previously difficult to access. We hope this will further the utilization of graph machine learning in neuroimaging.

## Data Availability

All data used in the study are available at https://www.humanconnectome.org/study/hcp-young-adult/overview (https://db.humanconnectome.org/data/HCP_1200) and http://www.dutchconnectomelab.nl/10Kdata/. All code used in the analysis is publicly available (https://github.com/wwu-mmll/photonai_graph_usecases; https://github.com/wwu-mmll/photonai_graph).

https://db.humanconnectome.org/data/HCP_1200

https://www.humanconnectome.org/study/hcp-young-adult/overview

## ACKNOWLEDGEMENTS

Data were provided in part by the Human Connectome Project, MGH-USC Consortium (Principal Investigators: Bruce R. Rosen, Arthur W. Toga and Van Wedeen; U01MH093765) funded by the NIH Blueprint Initiative for Neuroscience Research grant; the National Institutes of Health grant P41EB015896; and the Instrumentation Grants S10RR023043, 1S10RR023401, 1S10RR019307.

## Supplementary Material

**Fig. 5.**
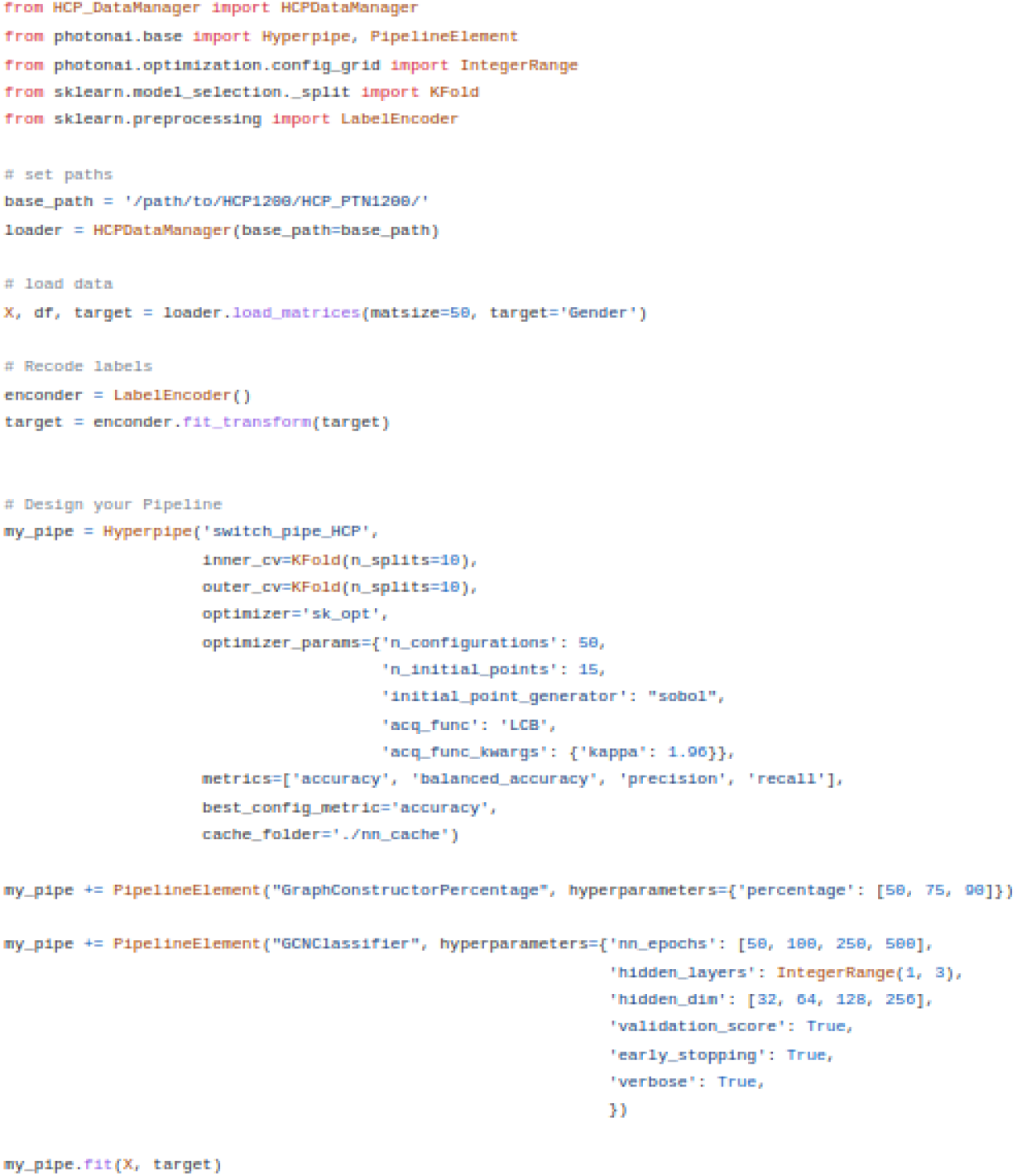
Prediction Script. The HCP-YA Sex Prediction pipeline can be built in 43 lines of code. Line 19-43 define and fit the pipeline, meaning that the entire pipeline optimization strategy, optimizer parameters, validation metrics, Graph Constructor, Graph Neural Network and pipeline fitting can be defined within 24 lines of code. This significantly decreases development time and increases accessibility.

